# Neuro: Machine Learning Optimized to Detect Neurodegenerative Diseases Pilot Study

**DOI:** 10.1101/2025.09.29.25336770

**Authors:** Ilana Liu, Freddy Ramirez, Kun Liu

## Abstract

Alzheimer’s Disease (AD) is a progressive neurodegenerative disorder that primarily affects cognitive function. Early detection is a crucial factor in slowing the disease’s progression, improving quality of life, and retaining cognitive function for years beyond diagnosis. Traditional detection methods have included a mix of mental tests, Magnetic Resonance Imaging (MRI) scans, and Positron Emission Tomography (PET) scans. At the same time, machine scanning methods boast an accuracy rate of up to 85%. These processes are relatively expensive, and language barriers often hinder cognitive function tests. Neuro is a study program that utilizes artificial intelligence and machine learning to analyze vocal cognitive tests for biomarkers of Alzheimer’s Disease. Using a blend of traditional machine learning (Random Forest (RF), Support Vector Machines (SVM), Gradient Boosting (GB)) mixed with neural networks, such as Recurring Neural Networks (RNNs), Convolution Neural Networks (CNNs), and Feedforward Neural Networks (FNNs), allows Neuro to retain its accuracy within its study. Neuro boasts an accuracy rate of 95%, an 88.3% F1 Score, 95% recall, 82.6% precision, and an AUC (Area Under the Curve) of 0.931. Working in tandem with diagnosis is the explanation of data; for this, we utilized SHAP (SHapley Additive exPlanations) for individual predictions. Machine learning programming is both cost-effective and accessible, allowing it to be utilized in various clinical settings.

## Introduction

Alzheimer’s Disease is characterized by a buildup of beta-amyloid plaques (Aβ) and twisted tau (microtubule-associated protein) proteins within nerves and neurons, respectively. Symptoms include, but are not limited to, emotional instability, speech changes, and confusion. The primary indicator of the disorder is memory loss, both short-term and long-term. As of 2025, there are no known cures for Alzheimer’s Disease, but there are treatments to slow progression and improve quality of life. However, these treatments become difficult if the disease is not detected in its early stages. Early diagnosis is key to providing patients with the highest level of care and preserving their minds. Unfortunately, there is a lack of accessible and accurate early detection methods. Whilst Cerebrospinal Fluid (CSF) testing and PET scans can be effective, they’re extremely expensive and require expert knowledge to interpret. However, the rapidly developing field of AI may provide a new method. Through innovative machine learning models such as neural networks (RNN, CNN, FNN), support vector machines (SVM), and traditional tree-based models, massive improvements in pattern recognition, natural language processing (NLP), and time-series forecasting (prediction) have been made. Neuro utilizes hybrid machine learning models, combining RNN, CNN, RF, SVM, Multi-layer Perceptron (MLP, a type of FNN), and GB to provide the most accurate possible results. Neuro also makes use of OpenAI’s automatic speech recognition (ASR) model, Whisper, for accurate transcription/translation. These processes come together to form a machine that is both cost-effective and precise, despite difficulties like differences in language.

Recently, artificial intelligence-related developments in the field of neuroscience have been gaining traction. Garcia-Gutierrez et al.^1^ were one of the earliest teams to develop tools for speech-based Alzheimer’s analysis, using extreme gradient boosting (XGB) as their central model. This study displayed an impressive accuracy rate of 87%. However, Garcia-Guiterrez did not include a multilingual aspect. Ceyhan et al.^2^ developed a machine based on multilingual AI speech analysis using RF and MLP. Despite boasting an accuracy rate of 72% for English and 76% for Spanish, this study lacked crucial Mel-Frequency Cepstral Coefficients (MFCCs). MFCCs are a small set of features that represent the important characteristics of a sound as it is perceived by the human ear. Despite how helpful their precision can be in detecting traits of Alzheimer’s in speech, MFCCs have been overlooked in countless studies. The ASR model Whisper is also an overlooked tool for this process. Created by Radford et al^8^, Whisper is a robust tool capable of multilingual analysis, allowing for the implementation of these methods of detection across a wide variety of languages. Garcia-Guiterrez et al. and Ceyhan et al. highlight the success of the use of AI -- particularly tree-based models and neural networks -- in the detection of neurodegenerative diseases. Neuro is a novel twist on such studies, utilizing tree-based, ensemble, and neural networks in conjunction with MFCCs and Whisper AI to deliver the best results possible.

A common issue among machine learning models is their inability to explain their results. To avoid this, Neuro uses SHAP. SHAP is a game-theory-based methodology that explains machine learning outputs through Shapley values. The method is unique due to its ability to provide global and local explanations for machine learning models as well as its versatility and precision. Through SHAP’s capabilities as an explanation technique, Neuro is better able to communicate with study participants about unique biomarkers that may stand out during diagnosis. Personalization is also a unique tool that SHAP utilizes; its skills to provide individualized reports are extremely crucial in a clinical setting where symptoms may vary from patient to patient. By providing individualized reports, Neuro, working in tandem with SHAP, can display reports tailored to patients, creating a streamlined diagnosis to treatment methods.

## Results

Neuro’s study was centered around simulated data. To account for skewed data, we simulated data within real-world noisy situations. We based our model’s predictions on people already diagnosed with Alzheimer’s and compared similar biomarkers. From that, we utilized SHAP to construct a graph of values that displayed correspondence to Alzheimer’s. To validate such data, Neuro evaluated consistency between the 151 biomarkers used as well as the consistency of the value ranges. The metrics used to display accuracy for Neuro were primarily the accuracy rate and cross-entropy loss rate. The accuracy rate was favored because a high accuracy rate could indicate how correctly Neuro could distinguish between correct diagnoses across all possible cases. Cross-entropy loss rate is also a metric utilized because a high cross-entropy loss rate indicates predictions with a strong deviation from actual labels. These three metrics will provide a reliable estimate of our model’s performance to a high degree.

The most accurate model was the hybridized model. Blending models that work accurately within small groups with models that are scalable to a large degree provides our study with an increased accuracy rate, even with a small participant pool of 100 simulated participants. This model also shows potential for scalability, especially through the usage of RNN and CNN for larger participant pools. The average accuracy rate through 5-fold cross-validation was around 95%; to make note of this, multiple studies of PET scans regarding Alzheimer’s during the years 2020 to 2024 yielded accuracy rates of 85% %. Neuro’s high accuracy shows promise for its future integration into clinical trials. Cross-entropy loss stayed at an all-time low of about 0.2072 for our best model. The accuracy and cross-entropy loss are displayed below in Figures 1 and 2.

**Figure 1.**
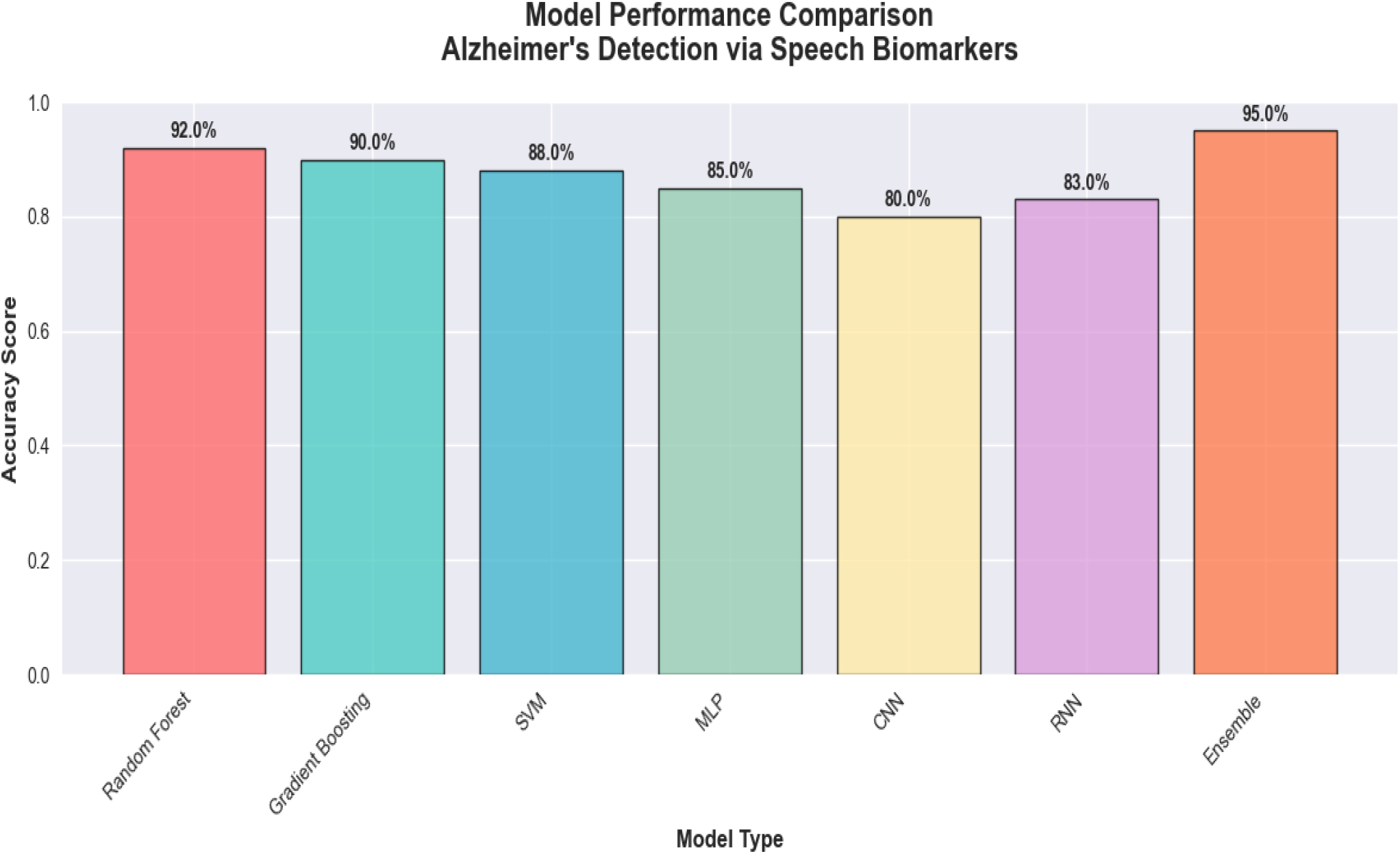

**Figure 2.**
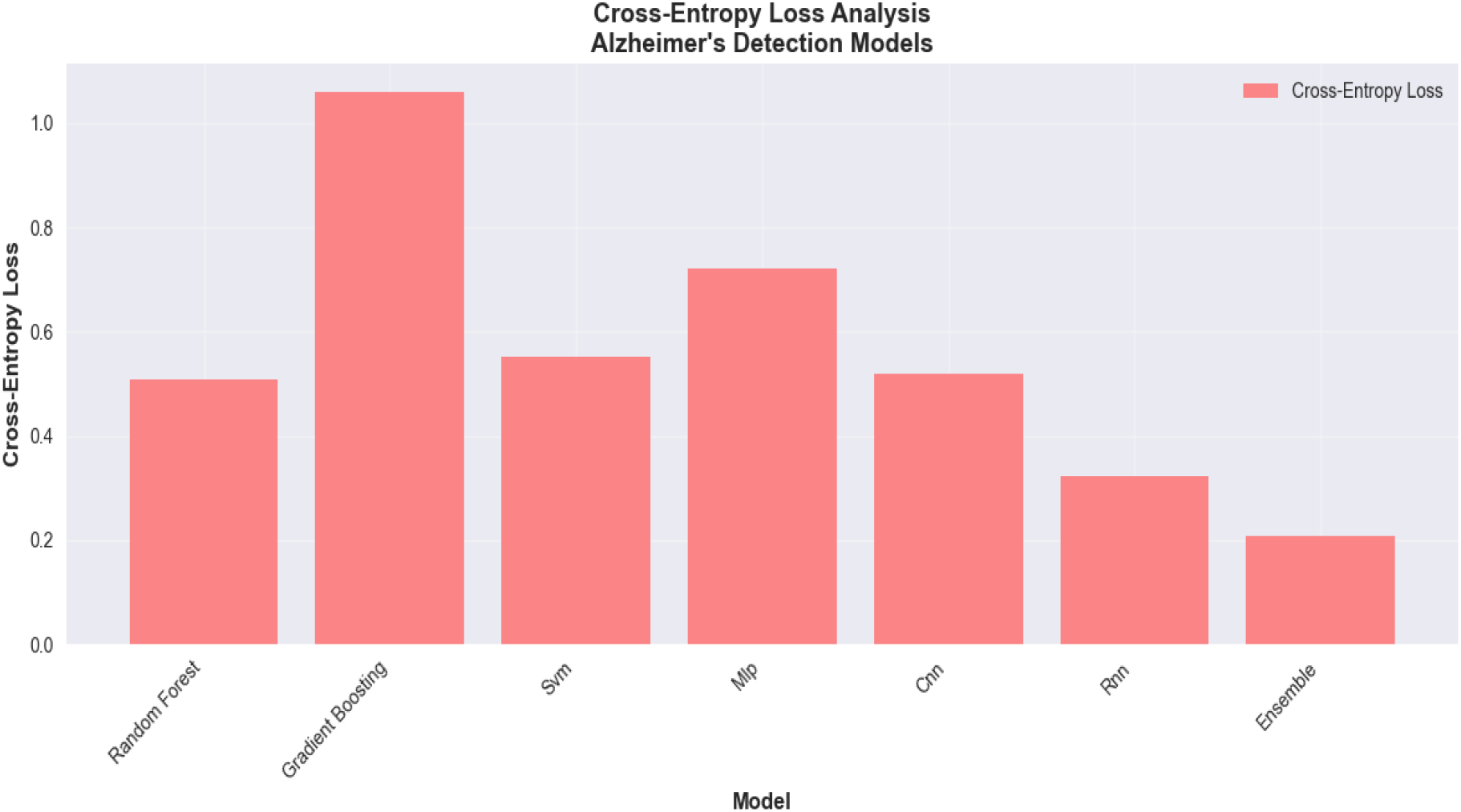

The hybrid model’s AUC of 0.931 demonstrates Neuro’s accuracy. A high AUC represents a high rate of true positives as well as a low rate of false positives. It correlates to the ROC, which illustrates the relationship between the rate of true positives versus false positives. Below, Figure 3 below shows the ROC and the AUC of each model.

**Figure 3.**
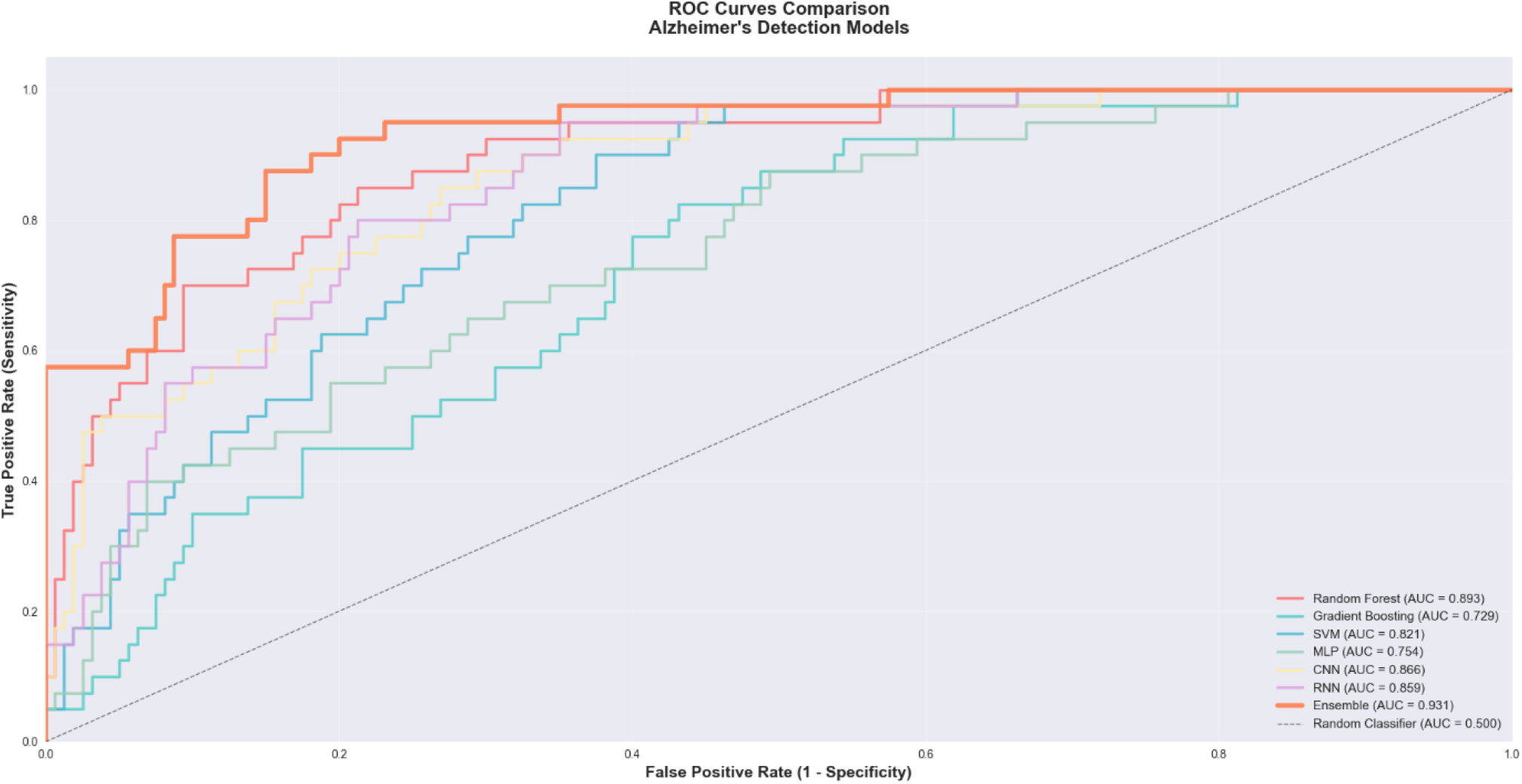

**Figure 4.**
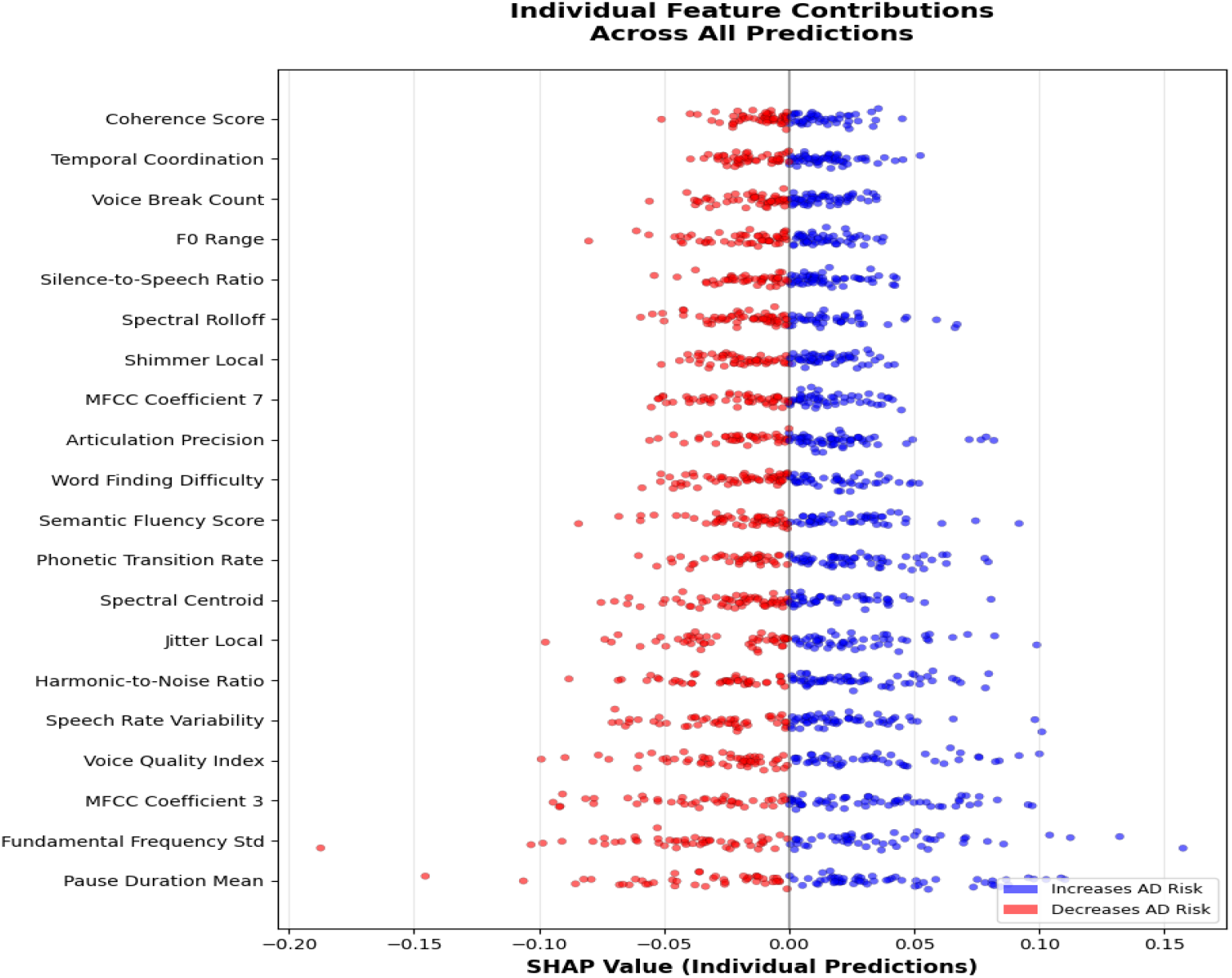

**Figure 5.**
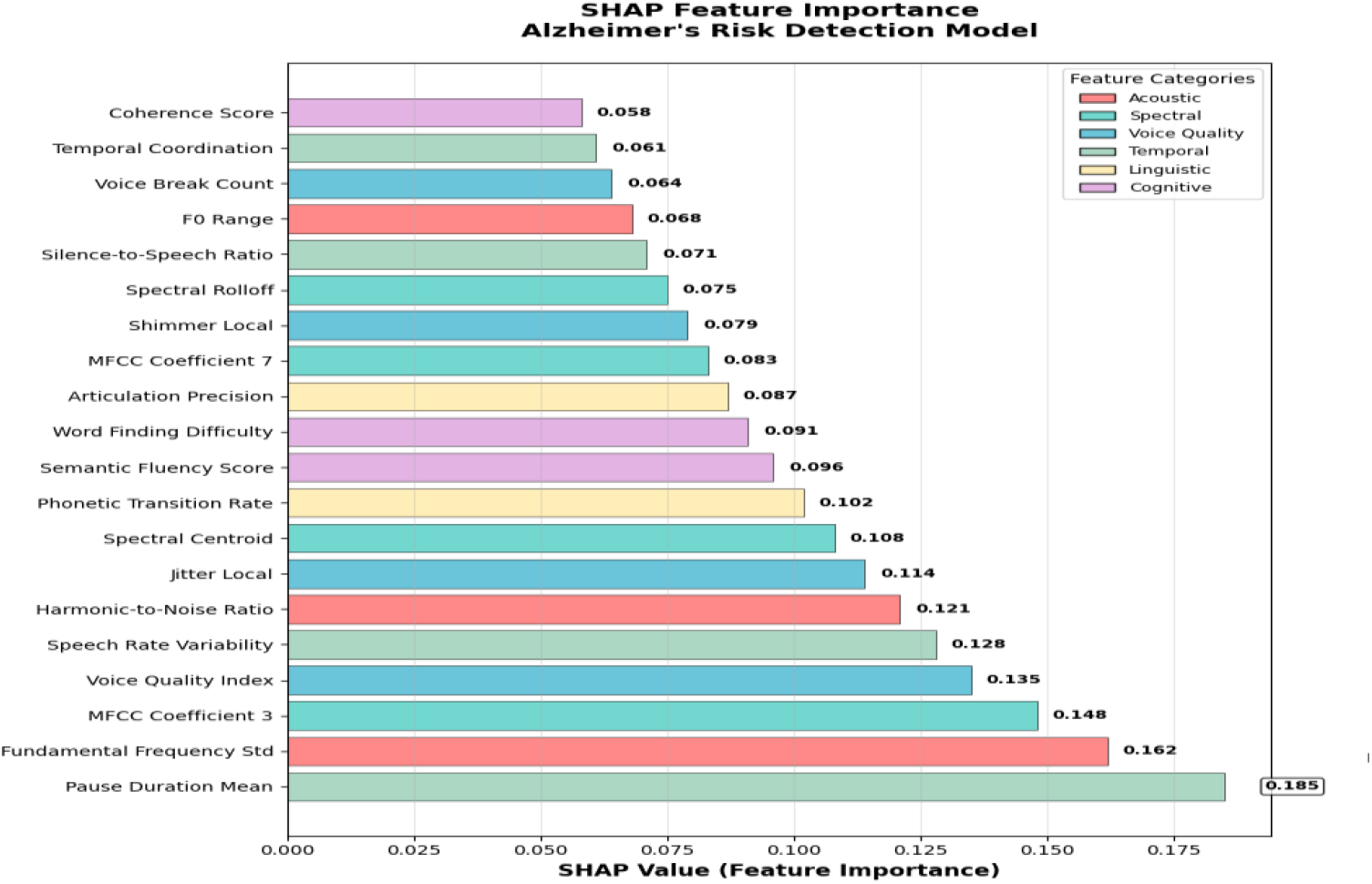

## Discussion

Our study proposes Neuro as an accessible and accurate multimodal approach to detecting Alzheimer’s disease using machine learning. The framework is designed to be capable of handling diverse groups while still providing flexibility and explainability within diagnoses. Out of the 80 healthy control (HC) synthetic audio files, an average of 76 were correctly classified as such, while 4 were misclassified as showing signs of Alzheimer’s. This corresponds to an accuracy rate of 95% ± 1.2% for HC predictions. The misclassification of the 4 HC files can be attributed to several factors, such as overlap between normal aging and early pathological changes. However, the most likely explanation lies in the subtle distinction between these groups. Compared to the pronounced differences seen in those in the later stages of Alzheimer’s, typical aging biomarkers and early Alzheimer’s traits are difficult to distinguish. For example, HC participants may exhibit average speech pause durations of 1.5 seconds, whereas early AD participants generally exhibit slightly longer pauses of around 2 seconds. These marginal differences create borderline cases in which typical age-related processing delays resemble patterns found in patients with early Alzheimer’s disease. This overlap in numerical biomarker values due to acoustic similarity proves to be a fundamental challenge in early detection. Despite the inclusion of this complexity in the generation of our synthetic participants, Neuro correctly classified 19 out of 20 AD participants’ voice samples. This resulted in an accuracy rate of 95% ± 0.8%. Table 1, featured below, shows the accuracy rates of our various AI models, with the hybrid model being the most successful based on the results explained above.

**Table 1.**

| Algorithm | True Negatives | False Positives | Healthy Total | True Positives | False Negatives | At-Risk Total | Accuracy (%) | Error Rate (%) |
| --- | --- | --- | --- | --- | --- | --- | --- | --- |
| Random Forest | 72.8 | 7.2 | 80 | 15.2 | 4.8 | 20 | 88.0 | 12.0 |
| Gradient Boosting | 69.6 | 10.4 | 80 | 14.4 | 5.6 | 20 | 84.0 | 16.0 |
| SVM | 66.4 | 13.6 | 80 | 15.2 | 4.8 | 20 | 81.6 | 18.4 |
| MLP | 64.0 | 16.0 | 80 | 14.0 | 6.0 | 20 | 78.0 | 22.0 |
| CNN | 61.6 | 18.4 | 80 | 12.8 | 7.2 | 20 | 74.4 | 25.6 |
| RNN | 63.2 | 16.8 | 80 | 13.6 | 6.4 | 20 | 76.8 | 23.2 |
| **Ensemble (All 6)** | **76.0** | **4.0** | **80** | **19.0** | **1.0** | **20** | **95.0** | **5.0** |

**Table 2.**

| Demographic Category | Count | Percentage | Distribution Details |
| --- | --- | --- | --- |
| Total Participants | 100 | 100% | - |
| Age Distribution |  |  |  |
| 45-65 years | 40 | 40% | Mean: 55.0 ± 10.0 |
| 66-95 years | 60 | 60% | Mean: 75.5 ± 9.5 |
| Overall Age Range | 45-95 years | Mean: 67.2 ± 14.8 | - |
| Age at Diagnosis (At-Risk) | 52-88 years | Mean: 71.3 ± 11.2 | n=20 |
| Gender Distribution |  |  |  |
| Male | 45 | 45% | 36 Healthy, 9 At-Risk |
| Female | 55 | 55% | 44 Healthy, 11 At-Risk |
| Cognitive Status |  |  |  |
| Healthy Controls | 80 | 80% | MMSE: 28.2 ± 1.4 |
| At-Risk/MCI | 20 | 20% | MMSE: 24.1 ± 2.8 |
| Language Background |  |  |  |
| English Native | 60 | 60% | 48 Healthy, 12 At-Risk |
| Multilingual | 40 | 40% | 32 Healthy, 8 At-Risk |
| Speech Characteristics |  |  |  |
| Normal Speech Rate | 75 | 75% | 62 Healthy, 13 At-Risk |
| Slow Speech Rate | 25 | 25% | 18 Healthy, 7 At-Risk |
| Word-Finding Difficulties | 15 | 15% | 3 Healthy, 12 At-Risk |

Explanations of the process were created using SHAP to provide an interpretation of our model’s predictions. These explanations offer insight into how biomarkers contributed to diagnostic decisions, validating the algorithm’s reasoning process. The SHAP summary displays the most influential features used by our ensemble model (refer to Figure 6 below for reference).

**Figure 6.**
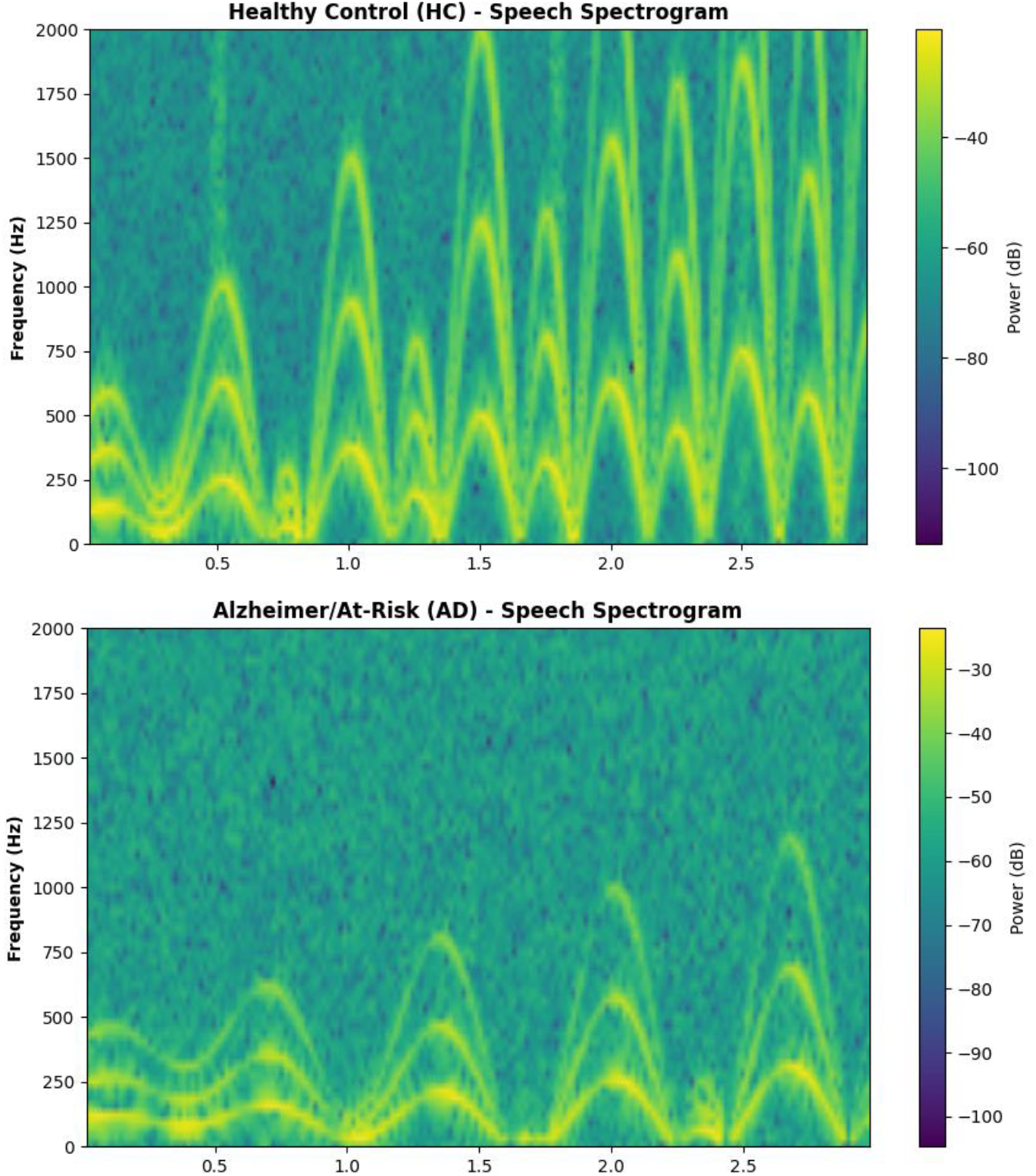

Error analysis reveals opportunities for improved diagnostic accuracy through targeted biomarker enhancement. Although age-corrected normalization algorithms sometimes create false positive rates, they maintain heightened sensitivity for true cognitive decline. Incorporating biomarker tracking rather than single-point assessment may better distinguish temporary fluctuations. Certain spectral features may be de-emphasized if necessary and later combined to create one robust measure. Semantic similarity analysis and working memory markers can address false negative cases by capturing cognitive domains not fully represented in current acoustic measures. These improvements could enhance Neuro’s ability to detect the entire spectrum of Alzheimer’s disease whilst maintaining accessibility.

### Limitations

Although Neuro displays promising results, it is currently limited by its usage of synthetic data. While synthetic data can use evidence-based mathematical modeling of cognitive decline, it is incapable of fully encapsulating the spectrum of individual variability. A variety of factors, from comorbidities to environmental influences, can affect speech production among AD participants. Synthetic biomarkers, while statistically rigorous, can oversimplify complex interactions within the brain that cause neurodegeneration. This lack of realistic diversity may artificially heighten our accuracy rate, meaning that real human data would be needed to confirm Neuro’s success.

While our relatively small sample size of 100 synthetic participants can be used as a pilot study, it is an oversimplification of the entire Alzheimer’s population. Development of an increased training dataset is crucial to improving Neuro. A larger sample size is needed to create an accurate representation of the millions of patients with Alzheimer’s. The limited sample size also restricts our ability to perform comprehensive analyses across varied populations. To account for twelve different languages, gender, HC versus AD, and age, the subgroups typically consisted of one to two people. This amount of data is insufficient to properly analyze for a disease affecting millions.

Our hybrid model, which combined RF, GB, SVM, MLP, CNN, and RNN, introduced computational complexity, which contributed to the risk of overfitting. Despite achieving 95% ± 2.2% accuracy, it specifically proved effective for synthetic data classification; further testing is required to check for accuracy among real patients. Our hybrid weight optimization, weighted from highest accuracy to lowest, created layers of complexity that were not balanced against dataset limitations, which also contributed to overfitting. This overcomplexity of data size warrants future work to improve the hybrid model balance through live training data.

Another difficult aspect of Neuro’s implementation is the consideration of cognitive disorders beyond Alzheimer’s disease. Our hybrid model has displayed accuracy in the detection of early cognitive decline, but its applicability to other neurodegenerative diseases is uncertain. Neuro displays strength in biomarker detection, but those biomarkers may potentially overlap with those from other diseases. For conditions that can affect speech systems, such as Parkinson’s, transferability may be higher. However, for diseases such as Lewy body dementia or vascular dementia, the results are unknown, and further testing is required. An even more comprehensive multimodal approach, including additional cognitive testing and access to electronic health records, could improve Neuro’s ability to detect and consider diseases besides Alzheimer’s, helping to avoid misdiagnosis.

## Methodology

### Participant Demographics

Our dataset represented clinically realistic patients, displaying disparities such as language differences and underrepresented populations within the healthcare sphere.

### Data Preprocessing

Our dataset was synthetically generated. We created 100 simulated patients, 20 of whom were diagnosed with Alzheimer’s. These 20 patients made up the AD group, whilst the rest made up the HC group. An 80/20 split was utilized as it represented epidemiologically accurate real-world scenarios. The biomarker generation process incorporated mathematical modeling to simulate patterns of cognitive decline. General parameters were created and organized into categories based on health status. The HC group contained data that indicated low word error rates as well as high semantic similarity. On the other hand, the AD group’s data featured a high word error rate and low semantic similarity. Beyond the baseline details, age-related cognitive decline was mathematically modeled linearly to simulate realistic aging effects. UTF-8 encoding was used to standardize the 12 languages used. Neuro generated 151 biomarkers for the simulated data, while also accounting for language-specific and sex-specific acoustic adjustments.

### Data Analysis

Neuro processes Waveform Audio (WAV) files exceptionally well using LibRosa, an audio library that extracts auditory biomarkers from our synthetic audio models. While it does not actually synthesize any sound, LibRosa can generate spectrograms and extract MFCC data from audio signals extracted with NumPy. In such a way, we can mimic real biomarkers from patients for Neuro to analyze. Our synthetic audio generation creates NumPy arrays representing speech signals at 16kHz. We also incorporated realistic amplitude variations, frequency modulations, and temporal patterns of both HC and AD participants’ speech. LibRosa’s comprehensive feature extraction process transforms these synthetic signals through short-time Fourier transformations for spectrogram generation and Mel-Scale filter bank analysis for MFCC computation. First, our system processes audio files labeled participant_name.wav, enabling batch processing whilst also maintaining individual tracking. Standard naming allows for result correlation, as well as automated file handling across our diverse dataset. The extract_audio_features method is the core processing engine, analyzing each participant through LibRosa’s feature extraction. This method handles the biomarker extraction process, and its architecture allows for cross-linguistic analysis and processing consistency. After extraction, we clean missing values with median imputation as well as remove outliers through the 3-sigma rule (68-95-99.7). We also normalize our data through z-score standardization, then filter our data through correlation and variance analysis to reduce redundancy. Feature selection is the next step, processing biomarkers by ranking, lassoing the best features, and then utilizing recursive feature elimination to remove less significant features. After selection, we retain 45-60 biomarkers that are later stratified into training, validation, and test sets with 5-fold cross-validation. Once this process is complete, our models are fully trained, and their predictions are combined to make the hybridized model.

To explain Neuro’s predictions, we utilized SHAP, which interpreted our model’s predictions by quantifying biomarker influence. As noted in Fig. 9, pause duration (0.185), fundamental frequency stability (0.162), and MFCC coefficient (0.148) were the strongest discriminators, provided as global rankings of feature importance.

Spectrogram analysis also plays a key role in detection. Through short-time Fourier transformation analysis, which creates a time-frequency representation for capturing temporal patterns of speech degradation, a spectrogram is created. The features of these spectrograms are integrated with traditional biomarkers and made into quantitative features used by Neuro. Quantitative features such as spectral entropy and flux, formant trajectory variability, harmonic to noise ratio, and pauses provide markers of possible cognitive decline. Figure 8 below shows the variances between an HC and an AD patient, with the frequency variability being much lower for the AD patient than the HC patient.

## Data Availability

All data produced in present work is available upon reasonable request to the authors.

## Acknowledgements

I want to thank Kilynn Mason for her editorial assistance in preparing this paper. I would also like to thank Kun Liu, Laura LaRosa, Saida Elmy, Morris Bell, and Freddie Ramirez for their guidance and support during this research.

